# International estimates of intended uptake and refusal of COVID-19 vaccines: A rapid systematic review and meta-analysis of large nationally representative samples

**DOI:** 10.1101/2020.12.01.20241729

**Authors:** Eric Robinson, Andrew Jones, India Lesser, Michael Daly

## Abstract

**Background:** Widespread uptake of COVID-19 vaccines will be essential to extinguishing the COVID-19 pandemic. Vaccines have been developed in unprecedented time and hesitancy towards vaccination among the general population is unclear.

**Methods:** Systematic review and meta-analysis of studies using large nationally representative samples (n≥1000) to examine the percentage of the population intending to vaccinate, unsure, or intending to refuse a COVID-19 vaccine when available. Generic inverse meta-analysis and meta-regression were used to pool estimates and examine time trends. PubMed, Scopus and pre-printer servers were searched from January-November, 2020. Registered on PROSPERO (CRD42020223132).

**Findings:** Twenty-eight nationally representative samples (n = 58,656) from 13 countries indicate that as the pandemic has progressed, the percentage of people intending to vaccinate and refuse vaccination have been decreasing and increasing respectively. Pooled data from surveys conducted during June-October suggest that 60% (95% CI: 49% to 69%) intend to vaccinate and 20% (95% CI: 13% to 29%) intend to refuse vaccination, although intentions vary substantially between samples and countries (I^2^ > 90%). Being female, younger, of lower income or education level and belonging to an ethnic minority group were consistently associated with being less likely to intend to vaccinate. Findings were consistent across higher vs. lower quality studies.

**Interpretation:** Intentions to be vaccinated when a COVID-19 vaccine becomes available have been declining globally and there is an urgent need to address social inequalities in vaccine hesitancy and promote widespread uptake of vaccines as they become available.

**Funding:** N/A

**Research in context:** *Evidence before this study:* We searched PubMed, Scopus and pre-print servers for manuscripts from January to November, 2020, reporting on studies examining intentions to be vaccinated against COVID-19 in large nationally representative samples (N≥1000). No language restrictions were applied. Search terms were [(COVID OR coronavirus OR SARS-COV-2) AND (Vaccine OR Vaccination) AND (Inten* OR willing* OR attitud* OR hypothetical)]. From 792 articles, we identified 20 eligible articles reporting on 28 nationally representative samples.

*Added value of this study:* This is the first systematic study and meta-analysis to estimate the proportion of the global population willing to be vaccinated against vs. intending to refuse a vaccine when COVID-19 vaccines become available and how this trend has changed over time, using large and nationally representative samples. Results indicate that COVID-19 vaccination intentions vary substantially across countries, the percentage of the population intending to be vaccinated has declined across countries as the pandemic has progressed (March-May estimate: 79%, June-October estimate: 60%) and a growing number report intending to refuse a vaccine, when available (March-May estimate: 12%, June-October estimate: 20%). There is consistent socio-demographic patterning of vaccination intentions; being female, younger, of lower income or education level and belonging to an ethnic minority group are associated with a reduced likelihood of intending to be vaccinated when a vaccine become available.

*Implications of all the available evidence:* Intentions to vaccinate against COVID-19 among the general public when a vaccine becomes available have been declining and this will limit the effectiveness of COVID-19 vaccination programmes. Findings highlight the need to improve public acceptability, trust and concern over the safety and benefit of COVID-19 vaccines and target vaccine uptake in disadvantaged groups who have already been disproportionately affected by the pandemic.

## Introduction

The COVID-19 pandemic has resulted in more than 1 million deaths worldwide from March to October, 2020^1^ and is likely to continue to have far reaching impacts on healthcare systems ^2^. The development of vaccines against COVID-19 has been occurring at unprecedented speed, and as of November, 2020, there are multiple candidate vaccines in the final stages of testing^3^. The success of any vaccination programme is dependent on the proportion of the population willing to be vaccinated and based on recent estimates it is likely that up to three quarters of the population may require vaccination to bring an end to the pandemic^4,5^.

Early in the pandemic a small number of studies surveyed adults to gauge public willingness to be vaccinated against COVID-19 and although a number of studies were reliant on non-representative convenience samples, the majority of the populations sampled intended to vaccinate^6-9^. For example, a cross country survey found a relatively high average level of intended vaccination (72%), although sample sizes of individual countries were low (N = ∼600-800) and may not provide accurate nationally representative coverage^7^.

However, as the pandemic has evolved there have been reports of widespread misinformation about COVID-19^10^, distrust in government^11^ and public concerns about the safety of COVID-19 vaccines given their rapid development^12^, all of which may have affected vaccine uptake. It is also unclear whether vaccine acceptability will be socio-demographically patterned. Disadvantaged minority groups have previously been shown to be less likely to intend to be vaccinated for influenza^13,14^, although a systematic review concluded that other demographic patterning of previous influenza vaccination programmes is inconsistent^15^. Given that current evidence on socio-demographic patterning of COVID-19 vaccination intentions is lacking, it will be important to understand how vaccination intentions differ within and across countries to inform measures to improve public acceptability and uptake of vaccination programmes.

We conducted a rapid systematic review and meta-analysis of large nationally representative study samples to address the current lack of consensus on; i) the proportion of the population willing to be vaccinated against COVID-19 when a vaccine become available and how this differs across countries, ii) whether vaccination intentions have declined as the pandemic has progressed, iii) socio-demographic inequalities in intended vaccine uptake.

## Method

To inform mass COVID-19 vaccination programmes, we used rapid systematic review methodology^16^. Rapid reviews provide timely evidence synthesises whilst maintaining the rigour of traditional systematic reviews^17^ using expedited review processes ^17,18^, such as limiting databases searched or number of reviewers (e.g. cross-checking of a proportion of extraction as opposed to independent extraction by a second author).

### Eligibility criteria

We included studies that measured intentions to be vaccinated against coronavirus in nationally representative samples of the general public. Published journal articles and pre-prints were eligible for inclusion. News articles reporting on opinion polls (with no corresponding scientific report including methodology used) were not eligible. The review is registered on PROSPERO (CRD42020223132).

### Populations and study design

To be eligible studies were required to have used a sampling approach designed to be nationally representative on key population demographics of the country (e.g. gender, education level), such as quota sampling or random probability sampling. Sampling error is problematic when examining prevalence estimates in small sample sizes^19^. Because sampling error tends to be minimal at sample sizes of ≥1000^19^, as is considered common practice for nationally representative surveys^20^, we limited eligibility to studies with a sample of n≥1000 from the same country. Studies that collected non-general public samples (e.g. healthcare professionals, parents, students) were not eligible. Studies that used non-representative sampling (e.g. convenience sampling, snowball sampling) were not eligible and studies that lacked sufficient methodological information to determine sampling used were ineligible.

### Interest (measure)

To be eligible studies were required to include a question that measured intentions/willingness to use a vaccine for COVID-19 when one becomes available (e.g. ‘I would use a vaccine for COVID-19 when it becomes available’). Studies that exposed participants to information designed to alter vaccine intentions (e.g. experiments comparing how public health messages may improve vaccine intentions) were not eligible. Studies that compared willingness to take different types of vaccine (e.g. varying hypothetical effectiveness/price) were ineligible, unless they also included a questionnaire item measuring general willingness to take a COVID-19 vaccine.

### Outcome

Studies were required to report the proportion of participants responding to the different response options for the vaccine intention question (e.g. Yes vs. No, Willing vs. Unsure Vs. Unwilling, Likely vs. Undecided vs. Unlikely).

### Article identification strategy

During November 2020, we searched PUBMED and Scopus (2020 onwards) for published articles in peer reviewed journals. We used the following search terms: (COVID OR coronavirus OR SARS-COV-2) AND (Vaccine OR Vaccination) AND (Inten* OR willing* OR attitud* OR hypothetical). We also searched three pre-print servers; Open Science Framework (which includes 30 other preprint archives, including PsychArxiv), MedrXiv and the Social Science Research Network (SSRN). One author conducted the initial title and abstract screening to exclude unrelated articles and a second author checked 25% of this (there were no discrepancies). A single author conducted full-text screening to determine eligibility and a second author cross-checked all eligibility decisions. For all eligible articles identified through searches, we used forward citation tracking (Google Scholar) and our knowledge of existing research to identify any further articles.

### Data extraction

For each study one author extracted information, all extraction was cross-checked by a second author and disagreements were resolved through discussion. We extracted the following information; bibliographic information, country, sampling procedure (e.g. quota vs. probability), sample size, month of survey, measure of vaccination intentions and results. We extracted results based on response options, resulting in % choosing response options indicative of definitely or probably yes, % definitive or probable no, % unsure (if measure allowed for the latter). If studies had multiple waves of data collection, we extracted results from the most recent wave. We also extracted information on whether studies reported results of analyses examining demographic predictors of vaccination intentions that were commonly examined (i.e. at least 5 studies) in studies. To be eligible for extraction, demographic predictors were required to be examined adjusting for other demographics (i.e. zero-order correlations were not eligible) in order to be confident of independent effects. We prioritised extraction of results of analyses that examined vaccine intentions (i.e. reference of category of yes vs. other (unsure/no combined). If this analysis was not available, in order of priority we favoured extracting demographic results for analyses examining yes vs. no, then yes vs. unsure.

### Risk of bias indicators

We considered quota-based samples as being higher in risk of bias than probability-based sampling, as the latter tends to be a more accurate/representative sampling approach^21,22^. We also reasoned that studies not having yet undergone formal peer review (pre-prints) may increase risk of bias and coded for this. We reasoned that studies that did not use an ‘unsure’ or ‘undecided’ response option (e.g. responses grouped into yes vs. no, as opposed to yes vs. unsure vs. no) may affect vaccine acceptance and rejection estimates, so we examined if pooled estimates of intended vaccination and intended refusal differed between these two types of study. For studies examining demographic predictors, we considered relatively smaller sample sizes (n<2500) as higher in risk of bias due to concerns over small numbers of cases in analyses when examining sub-groups. For example, with n ≥ 2500, for minority sub-groups (e.g. ∼2% of the population, such as Black people in the UK) there would be expected to be a minimum of 50 cases in analyses, as opposed to only 20 cases with n=1000. Studies of demographic predictors that adjusted for attitudinal predictors (e.g. attitudes towards vaccination) of vaccine intentions were considered higher in risk of bias, as inclusion may mask associations between demographics and vaccination intentions.

### Synthesis of evidence

We meta-analysed proportions of the samples ((Total sample N / 100) * % reporting) reporting: i) intending to vaccinate, ii) unsure, and iii) not intending to vaccinate. Analysis was performed using the ‘metafor’ package in R. We used a logit transformation on raw proportions in random effects, generic inverse variance meta-analyses with a restricted maximum-likelihood estimator. Transformations were conducted using the ‘escalc’ function in the metafor package. Back-transformed (inverse) logit values are presented in the text, whilst raw proportion data is presented in forest and funnel plots to aid interpretation. Heterogeneity was assessed with the I^2^ statistic. We conducted meta-regressions to examine the relationship between outcomes and month of study data collection (treating month as a continuous variable, e.g. March = 1, April = 2). We conducted leave-one-out analysis to examine the stability of the pooled estimates and identify any influential samples in the main analyses and meta-regressions. We conducted sub-group analyses to examine whether results differed between studies using quota vs. probability sampling and pre-prints vs. journal articles. We limited these analyses to studies which collected data early in the pandemic (March-May) to account for the majority of probability samples (3/4) and journal article sample (12/12) studies being conducted in this period. Sub-group analysis compared studies that did vs. did not allow for an ‘unsure’ response option. We also examined potential publication bias and small study effects (see online supplementary materials). Demographic predictors were measured and/or analysed differently across studies, so for each demographic we reported the proportion of studies finding evidence vs. no evidence of significant (p < .05) relationships with vaccination intentions, and whether results were similar when limited to studies that had larger sample sizes and did not adjust for attitudinal predictors in analyses.

## Results

### Study selection

A total of 792 unique articles were found through database searches and other sources. Of the 145 articles full-text screened, 20 articles reporting on 28 samples were eligible for inclusion. See Figure 1. For bibliographic information of all included articles, see online supplementary material.

**Figure 1.**
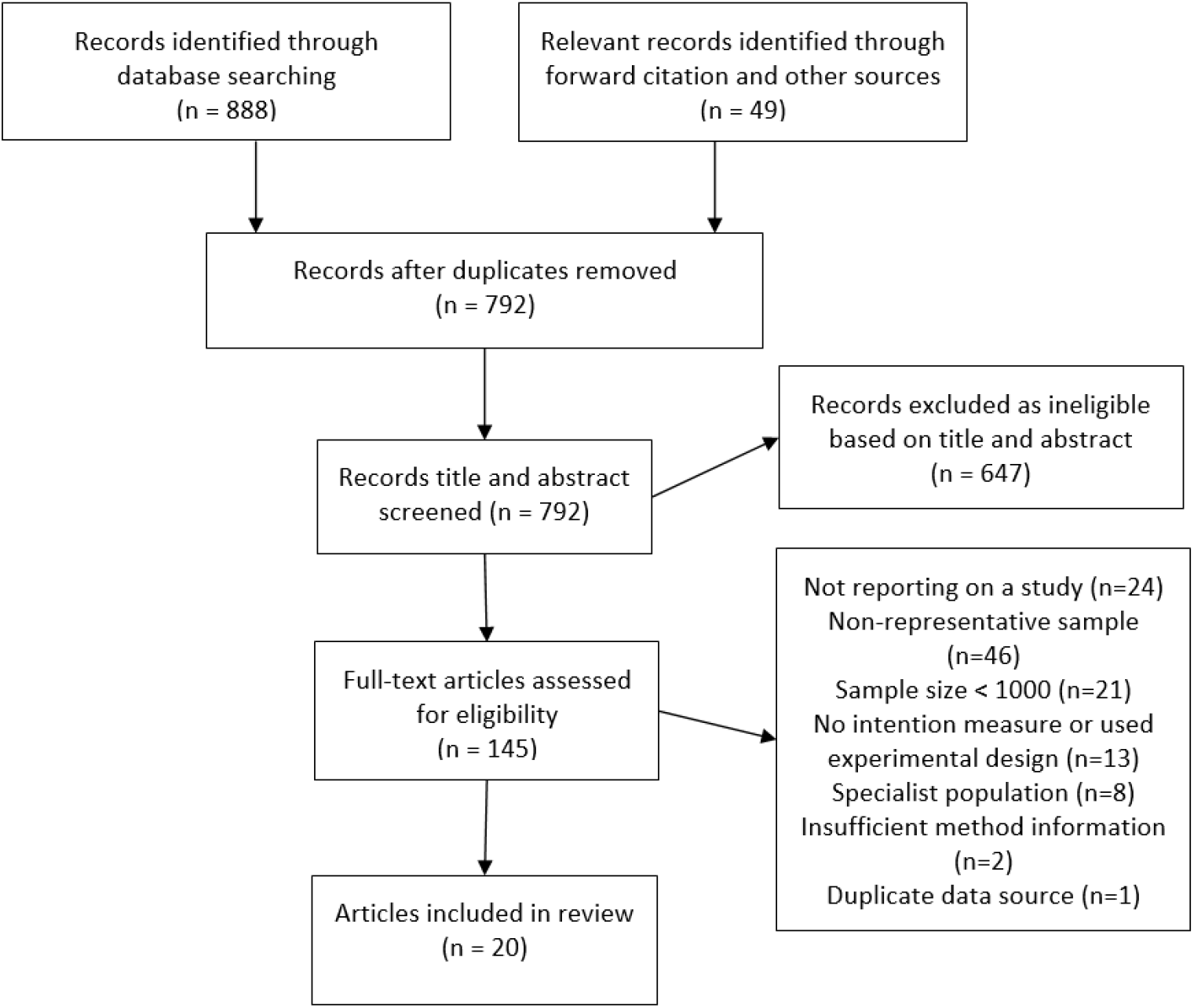
Study search and selection flowchart.

### Overview

Of the 28 samples included, the majority were from the UK and North America (7=US, 2=Canada, 7=UK) as well as samples from France (n=3), Australia, China, Denmark, Germany, Italy, Ireland Netherlands, Poland and Portugal. Full study information is reported in Table 1. Sample sizes ranged from 1,000 to 7,547 (median = 1198). Samples were collected in the early phases of the pandemic (March-May, n = 18) and later (June onwards, n = 10). Twenty-four samples used quota-based sampling and four used probability sampling, 12 articles were in peer-reviewed journals and 16 were pre-prints (at the time of identification).

**Table 1.** Study and sample information of eligible articles included in review

| Author | Country | Journal article or pre-print | Probability vs. quota sampling<sup>1</sup> | N | Month | Measure<sup>2</sup> | Intention results<sup>3</sup> |
| --- | --- | --- | --- | --- | --- | --- | --- |
| Edwards | Australia | Pre-print | Quota | 3,061 | August | If a safe and effective vaccine for COVID-19 is developed, would you... *Would definitely [Y], probably [Y], probably not [N], definitely not [N]* | Yes: 87%<br>No: 13% |
| Leigh | Canada | Pre-print | Quota | 1,996 | April-May | I will get vaccinated for the virus when it is developed... *Strongly disagree [N], disagree [N], somewhat disagree [N], undecided [U], somewhat agree [Y], agree [Y], strongly agree [Y]* | Yes: 76%<br>Unsure: 13%<br>No: 11% |
| Taylor | Canada | Journal article | Quota | 1,902 | May | If a vaccine for COVID-19 was available, would you get vaccinated? *Yes [Y], no [N]* | Yes: 80%<br>No: 20% |
| Wang | China | Journal article | Quota | 2,058 | March | Accept vaccination if the COVID-19 vaccine is successfully developed and approved for listing in the future? *Yes [Y], no [N]* | Yes: 91%<br>No: 9% |
| Neuman Bohme | Denmark | Journal article | Quota | 1,000 | April | Would you be willing to get vaccinated against the novel coronavirus?<br>*Yes [Y], no [N], not sure [U]* | Yes: 80%<br>Unsure: 12%<br>No: 8% |
| Neuman Bohme | France | Journal article | Quota | 1,000 | April | Would you be willing to get vaccinated against the novel coronavirus?<br>*Yes [Y], no [N], not sure [U]* | Yes: 62%<br>Unsure: 28%<br>No: 10% |
| Ward | France | Pre-print | Quota | 5,018 | April | Respondents were asked whether they would agree to get vaccinated if a vaccine against the COVID-19 was available:<br>*Certainly [Y], probably [Y], probably not [N], certainly not [N]* | Yes: 76%<br>No: 24% |
| Hacquain | France | Pre-print | Quota | 1,003 | September | Respondents were asked whether they would agree to get vaccinated if a vaccine against the COVID-19 were available<br>*Certainly [Y], probably [Y], probably not [N], certainly not [N]* | Yes: 52%<br>No: 48% |
| Neuman Bohme | Germany | Journal article | Quota | 1,002 | April | Would you be willing to get vaccinated against the novel coronavirus?<br>*Yes [Y], no [N], not sure [U]* | Yes: 70%<br>Unsure: 20%<br>No: 10% |
| Murphy | Ireland | Pre-print | Quota | 1,041 | March-April | If a new vaccine were to be developed that could prevent COVID-19, would you accept it for yourself?<br>*Yes [Y], maybe [Y], no [N]* | Yes: 91%<br>No: 10% |
| Graffigna | Italy | Journal article | Probability | 1,004 | May | Willingness to vaccinate against COVID-19 whenever the vaccine is available<br>*Absolutely likely [Y], very likely [Y], neither likely or unlikely [U], unlikely [N], not likely at all [N]* | Yes: 59%<br>Unsure: 26%<br>No: 15% |
| Neuman Bohme | Netherlands | Journal article | Quota | 1,012 | April | Would you be willing to get vaccinated against the novel coronavirus?<br>*Yes [Y], no [N], not sure [U]* | Yes: 73%<br>Unsure: 19%<br>No: 8% |
| Feleszko | Poland | Pre-print | Quota | 1,066 | June | If a vaccine against COVID-19 is available do you plan to vaccinate?<br>*Yes [Y], no [N], I do not know/it is difficult to answer [U]* | Yes: 37%<br>Unsure: 34%<br>No: 28% |
| Neuman Bohme | Portugal | Journal article | Quota | 1,064 | April | Would you be willing to get vaccinated against the novel coronavirus?<br>*Yes [Y], no [N], not sure [U]* | Yes: 75%<br>Unsure: 21%<br>No: 5% |
| Murphy | UK | Pre-print | Quota | 2,025 | March | If a new vaccine were to be developed that could prevent COVID-19, would you accept it for yourself?<br>*Yes [Y], maybe [Y], no [N]* | Yes: 94%<br>No: 6% |
| Neuman Bohme | UK | Journal article | Quota | 1,009 | April | Would you be willing to get vaccinated against the novel coronavirus?<br>*Yes [Y], no [N], not sure [U]* | Yes: 79%<br>Unsure: 15%<br>No: 6% |
| Freeman | UK (England) | Journal article | Quota | 2,501 | May | How likely it was that you would accept a vaccination for Coronavirus? *Definitely [Y], probably [Y], possibly [Y], probably not [N], definitely not [N]* | Yes: 88%<br>No: 12% |
| Roozenbeek | UK | Journal article | Quota | 1,150 | May | Participants were asked whether they would get vaccinated against COVID-19 if a vaccine were to become available *Yes [Y], no [N]* | Yes: 79%<br>No: 21% |
| McAndrew | UK | Pre-print | Quota | 1,663 | June | When a Coronavirus (COVID-19) vaccine becomes available, do you think you will or will not get vaccinated?<br>*Definitely will get vaccinated [Y], probably will get vaccinated [Y], probably will not get vaccinated [N], definitely will not get vaccinated [N], unsure [U]* | Yes: 69%<br>Unsure: 15%<br>No: 16% |
| Sherman | UK | Pre-print | Quota | 1,504 | July | How likely would you be to have a COVID-19 vaccination when a coronavirus vaccination becomes available? *Eleven-point scale from extremely unlikely to extremely likely. 0-2 [N], 3-7 [U], 8-10 [Y]* | Yes: 64%<br>Unsure: 27%<br>No: 9% |
| Loomba | UK | Pre-print | Quota | 4,001 | September | If a new coronavirus (COVID-19) vaccine became available, would you accept the vaccine for yourself? *Yes [Y], definitely [Y],* | Yes: 54%<br>Unsure: 40% |
|  |  |  |  |  |  | *unsure but leaning towards yes [U], unsure but leaning towards no [U], no definitely not [N]* | No:6% |
| Romer | US | Journal article | Probability | 1,050 | March | If there were a vaccine that protected you from getting the coronavirus, how likely, if at all, would you be to decide to be vaccinated? *Not at all likely [N], not too likely [N], somewhat likely [Y], very likely [Y]* | Yes: 86%<br>No: 15% |
| Taylor | US | Journal article | Quota | 1,772 | May | If a vaccine for COVID-19 was available, would you get vaccinated? *Yes [Y], no [N]* | Yes: 75%<br>No: 25% |
| Carpiano | US | Pre-print | Probability | 1,000 | May | If a vaccine against the coronavirus becomes available, do you plan to get vaccinated, or not? *Yes [Y], no [N], not sure [U]* | Yes: 50%<br>Unsure: 30%<br>No: 20% |
| Callaghan | US | Pre-print | Quota | 5,009 | May-June | Scientists around the world are working on developing a vaccine to protect individuals against the coronavirus. If a Vaccine is developed, would you pursue getting vaccinated for the coronavirus? *Yes [Y], no [N]* | Yes: 69%<br>No: 31% |
| McAndrew | US | Pre-print | Quota | 1,198 | June | When a Coronavirus (COVID-19) vaccine becomes available, do you think you will or will not get vaccinated? *Definitely will get vaccinated [Y], probably will get vaccinated [Y], probably will not get vaccinated [N], definitely will not get vaccinated [N], unsure [U]* | Yes: 59%<br>Unsure: 15%<br>No: 25% |
| Loomba | US | Pre-print | Quota | 4,000 | September | If a new coronavirus (COVID-19) vaccine became available, would you accept the vaccine for yourself? *Yes definitely [Y], unsure but leaning towards yes [U], unsure but leaning towards no [U], no definitely not [N]* | Yes: 44%<br>Unsure: 41%<br>No:15% |
| Daly | US | Pre-print | Probability | 7,547 | October | How likely are you to get vaccinated for coronavirus when a vaccine becomes available *Unsure [U], somewhat unlikely [N], very unlikely [N], somewhat likely [Y], very likely [Y]* | Yes: 54%<br>Unsure: 14%<br>No: 32% |

<sup>1</sup>If sampling method was unclear from the description in the study method, we confirmed use of quota vs. probability by searching for the data source online (e.g. panel provider website)
<sup>2</sup> [Y], [N] and [U] indicate extracted response options representing yes, no and unsure in the present meta-analysis
<sup>3</sup> Values may not equal 100 due to rounding of reported values in study manuscripts

### Intentions to vaccinate (Figures S1-S3)

Including all 28 samples collected from March-October, the pooled proportion reporting intending to vaccinate was .729 [n = 28, 95% CI: .666 to .784: I^2^ = 99.6%], the proportion reporting they would refuse a vaccine was .143 [n = 28 95% CI: .114 to .178: I^2^ = 99.3%] and the proportion reporting being unsure was .221 [n = 16, 95% CI: .178 to .271: I^2^ = 99.0%]. Values do not equal 100 as not all studies included an unsure response option. See supplementary materials document for results in full and Figure 2 for proportion of populations intending to vaccinate by country and month.

**Figure 2.**
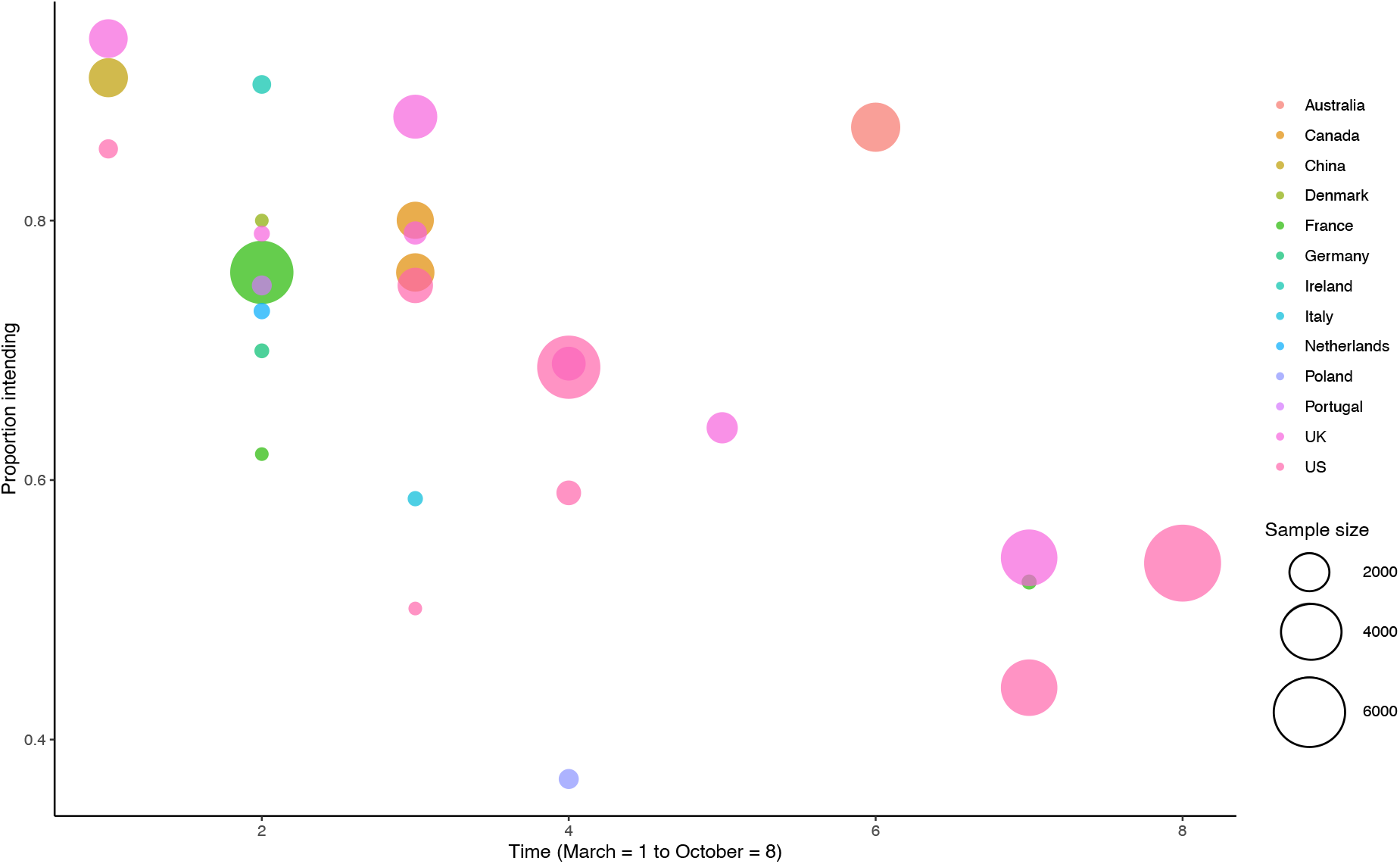
Proportion of populations intending to vaccinate by country and time.

### Presence vs. absence of ‘unsure’ response option in survey

There was a significant difference in the proportion intending to vaccinate when an ‘unsure’ response option was used vs. when there was no ‘unsure’ response option (X^2^(1) = 16.82, p < .001). When there was no unsure response option the proportion was .828 [95% CI: .759 to .880: I^2^ = 99.4%], and when unsure was a response option the proportion was .635 [95% CI: .569 to .670: I^2^ = 99.2%]. There was not a significant difference in the proportion intending to refuse a vaccine between samples with [.124, 95% CI: .092 to .163: I2 = 99.0%] vs. without [.172 [95% CI: .120 to .240: I^2^ = 99.4%] ‘unsure’ response options (X^2^(1) = 2.12, p = .146).

### Time trends

See Figures 2-4. There was a significant association between proportion of individuals intending to vaccinate and month study was conducted (coefficient = -.24 [95% CI: - .37 to .11], z = 3.55, p < .001) and also a significant association between intentions not to vaccinate and month of study (coefficient = .15 [95% CI: .03 to .28], z = 2.36, p = .018). Over time intentions to vaccinate decreased, and intentions not to vaccinate increased. For example, in studies (n = 18) that collected data during the early phase of the pandemic (March-May), the proportion intending to vaccinate was 79% and not intending to was 12%, compared with 60% and 20% June-October studies (n = 10). There was no association between proportion of individuals being unsure about vaccination and when the study was conducted (coefficient = .10 [95% CI: -.03 to .24], z = 1.54, p =.124]. Because samples varied by country, to confirm time trend findings were not explained by different countries being sampled earlier vs. later in the pandemic, we replicated the meta-regressions for the two countries with multiple samples collected during different months. Among UK samples (n = 7, coefficient = -.39 [95% CI: -.57 to -.21], z = 4.34, p < .001) and US samples (n = 7, coefficient = -.22 [95% CI: -.38 to -.05], z = 2.53, p = .014)] the same negative associations were observed as in the main analyses.

There was minimal evidence of publication bias and leave out one analyses indicated limited variation in estimates (online supplementary materials document). We found no evidence that results differed between samples reported in journal articles vs. pre-prints. There was minimal evidence of differences in findings between studies using probability vs. quota sampling, with the exception of ‘unsure’ responses being lower in quota samples (see online supplementary materials).

### Demographic predictors

Fourteen studies examined demographic predictors. See Table 2. In 12/14 studies older adults were significantly more likely to report intending to vaccinate than younger, one study found no effect of age and in one study young adults (<25 years) were more likely than middle aged adults, but not older adults. In 9/14 studies males were more likely to intend to vaccinate than females (no significant association in five studies). Higher education level was associated with intending to vaccinate in 7/14 studies (no association in seven). White ethnic groups were more likely to vaccinate in 7/11 studies (no association in four). Higher income was associated with intending to vaccinate in 8/9 studies and in one study there was no association. Presence of a health condition was examined in five studies and was non-significant in n=4 and not having a health condition was associated with intending to be vaccinated in one study. When analyses were limited to the five higher quality studies the role of demographic factors was more consistent; 5/5 studies found that older adults, males and higher education levels were associated with increased likelihood of intending to vaccinate. Similarly, 3/4 studies found that being white and 4/4 found that those on higher income were more likely to intend to vaccinate. Presence of a health condition was non-significant in n=2.

**Table 2.** Studies examining demographic predictors of vaccination intentions.

| **Author & Country** | **N<sup>1</sup>** | **Comparison information** | **Age** | **Gender** | **Education** | **Ethnicity** | **Physical health condition** | **Income** |
| --- | --- | --- | --- | --- | --- | --- | --- | --- |
| Edwards (Australia)* | 3,061 | Intention to vaccinate vs. other responses (do not intend and unsure) | Older adults more likely to vaccinate (55yrs+) | Males more likely to vaccinate | Higher education qualification more likely to vaccinate | Non-significant | Non-significant | Higher income more likely to vaccinate |
| Hacquin (France)* | 4,027 | Intention to vaccinate vs. do not intend | Older adults more likely to vaccinate (64yrs+) | Males more likely to vaccinate | Higher than high school qualification more likely to vaccinate | - | - | - |
| Ward (France) | 5,018 | Intention to vaccinate vs. do not intend (*Model included attitudinal measures*) | Older adults more likely to vaccinate (64yrs+) | Males more likely to vaccinate | Non-significant | - | - | Higher income more likely to vaccinate |
| Murphy (Ireland) | 1,041 | Intention to vaccinate vs. do not intend | Older adults (64yrs+) more likely to vaccinate than 35-44yr olds, but no other groups | Non-significant | Non-significant | Irish ethnicity more likely to vaccinate than non-Irish | No health condition more likely to vaccinate | Higher income more likely to vaccinate |
| Roozenbeek (UK) | 1,150 | Intention to vaccinate vs. do not intend (*Model included attitudinal measures*) | Older adults more likely to vaccinate | Non-significant | Non-significant | - | - | - |
| Loomba (UK)* | 4,001 | Intention to vaccinate vs. do not intend | Older adults (55yrs+) more likely to vaccinate than younger (<25 yrs) | Males more likely to vaccinate | Higher education qualification more likely to vaccinate | Whites more likely to vaccinate than Black and Asian (and other) | - | Higher income more likely to vaccinate |
| McAndrew (UK) | 1,663 | Intention to vaccinate vs. other responses (do not intend and unsure) | Older adults more likely to vaccinate | Non-significant | Non-significant | Non-significant | - | - |
| Murphy | 2,025 | Intention to vaccinate | Older adults (65yrs+) | Non-significant | Non-significant | Non-significant | Non- | Higher income |
| (UK) |  | vs. do not intend | more likely to vaccinate |  |  |  | significant | more likely to vaccinate |
| Sherman (UK) | 1,488 | Likelihood of being vaccinated, continuous measure | Older adults more likely to vaccinate | Non-significant | Non-significant | Non-significant | Non-significant | - |
| Daly (US)* | 7,547 | Intention to vaccinate vs. unsure | Older adults more likely to vaccinate (65yrs+) | Males more likely to vaccinate | Degree level education and above more likely to vaccinate | Whites more likely to vaccinate than African Americans (Black) | Non-significant | Higher income more likely to vaccinate |
| Carpiano (US) | 1,000 | Intention to vaccinate vs. unsure | Older adults (60yrs+) more likely to vaccinate than 30-59yrs | Males more likely to vaccinate | College level education and above more likely to vaccinate | Whites more likely to vaccinate than African Americans, Hispanic and other | - | Non-significant |
| Callaghan (US) | 5,009 | Intention to vaccinate vs. unsure (*Model included attitudinal measures*) | Older adults more likely to vaccinate | Males more likely to vaccinate | Non-significant | Whites more likely to vaccinate than African Americans | - | Higher income more likely to vaccinate |
| Loomba (US)* | 4,000 | Intention to vaccinate vs. do not intend | Young adults (<25yrs) more likely to vaccinate than other 25-54 | Males more likely to vaccinate | Higher educational qualification more likely to vaccinate | Whites more likely to vaccinate than African Americans (and other) | - | Higher income more likely to vaccinate |
| McAndrew (US) | 1,198 | Intention to vaccinate vs. other responses (do not intend and unsure) | Non-significant | Males more likely to vaccinate | College level education and above more likely to vaccinate | Whites more likely to vaccinate than African Americans | - | - |

<sup>1</sup> N may differ to Table 1 (e.g. Hacquin et al. examined demographic predictors across several waves of data collection, as opposed to only the final wave of data collection, as reported in Table 1)
\*indicates highest quality studies (N>2500, no inclusion of COVID attitudinal or previous vaccination behaviour variables in analyses)

## Discussion

Results of this systematic review and meta-analysis of 58,656 participants drawn from 28 large nationally representative study samples across 13 countries indicates that the percentage of the population intending to be vaccinated when a COVID-19 vaccine becomes available has declined markedly across countries as the pandemic has progressed. Numbers reporting that they will refuse a vaccine have increased over time and a substantial proportion of adults now intend to refuse a vaccine, when available (June-October estimate = 20%). There is also consistent socio-demographic patterning of vaccination intentions; being female, younger, of lower income or education level and belonging to an ethnic minority group are associated with a reduced likelihood of intending to be vaccinated when a vaccine become available.

Emerging evidence suggests that both exposure to misinformation about COVID-19 ^10,23^ and public concerns over the safety of vaccines^24^ may be contributing to the observed declines in intentions to be vaccinated, and this highlights the need for measures to address public acceptability, trust and concern over the safety and benefit of approved vaccines. As well as observing declines in intentions to vaccinate over time in our main analyses, we also note intentions differed markedly by country. When sampled during a similar period early on in the pandemic (March-April), 91% of adults in China reported intending to be vaccinated, compared to 76% of adults in France. However, the most recent estimate (September-October) in France is now 52% and similar to the US (54%).

Across studies there was very consistent socioeconomic patterning of vaccination intentions: lower income or education and ethnic minorities were less likely to intend to vaccinate. There was no evidence in any studies reviewed that presence of a chronic health condition was associated with increased vaccination intentions, even though these groups are at increased risk of dying from COVID-19^25^. Measures are required to maximise vaccine uptake in vulnerable and disadvantaged groups who have already been disproportionately affected by the pandemic, such as those from lower income and ethnic minority groups^25,26^. Strengths of the present research are that we limited evidence synthesis to study designs that allow for accurate estimates of population level intentions with minimal sampling error, as small studies of non-representative samples are likely to provide biased estimates. We found no evidence that the type of sampling method used had a meaningful impact on estimates and that studies reported in pre-prints produced similar effect estimates as peer-reviewed journals. Analyses also accounted for studies using different response formats and findings were similar.

We found evidence of declining vaccination intentions when data was analysed using meta-regression (continuous month-of-year variable), when comparing study estimates from early in the pandemic (March-May) to later (June-October) and when examining time trends within countries (UK, US). Two included individual studies (US, France) also reported vaccination intentions over time in the same population and results were consistent^24,27^. As included studies examined nationally representative samples we do not know whether a similar pattern of results would be expected among other population groups (e.g. healthcare workers) and research is required to address these questions^28,29^. Findings of the present research are limited to studies that met eligibility criteria for having used large and nationally representative sampling and this tended to be developed western countries.

## Conclusions

Intentions to vaccinate when a COVID-19 vaccine becomes available have been declining globally and there is an urgent need to address social inequalities in vaccine hesitancy and promote widespread uptake of vaccines as they become available.

### Data sharing

Study data files and analysis code are openly available on the Open Science Framework; https://osf.io/hj4ds/

## Contributors

The study was designed by ER. Data was collected by ER, AJ, IL and MD. AJ and ER analysed the data. The first draft was written by ER. All authors critically revised the manuscript and agree to be accountable for all aspects of the work.

## Acknowledgments

ER’s time was part-funded by the European Research Council and their support is gratefully acknowledged.

## Figure headings

**Figure S1.**
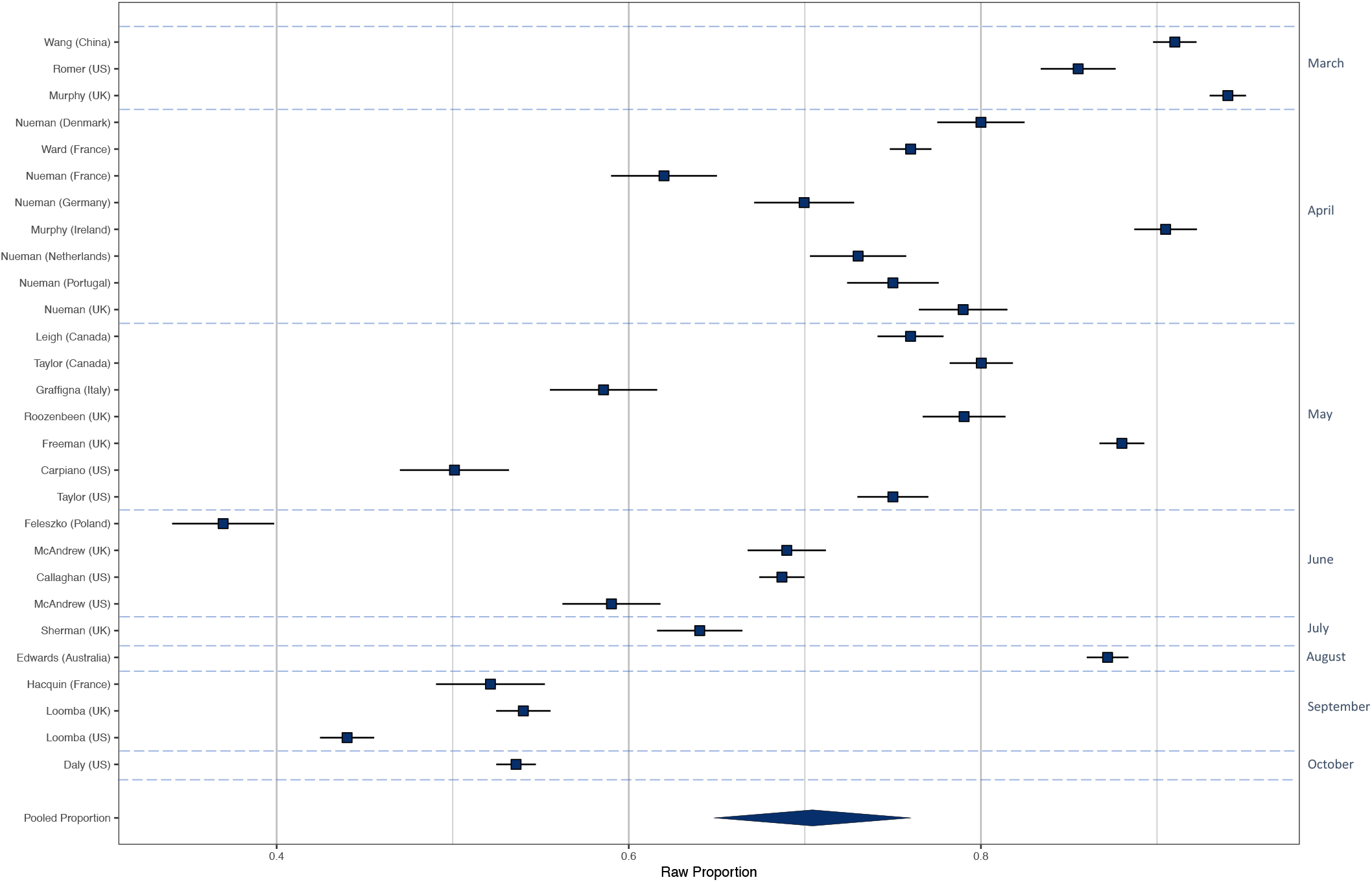
Raw proportions of intentions to vaccinate across the 28 samples.

**Figure S2.**
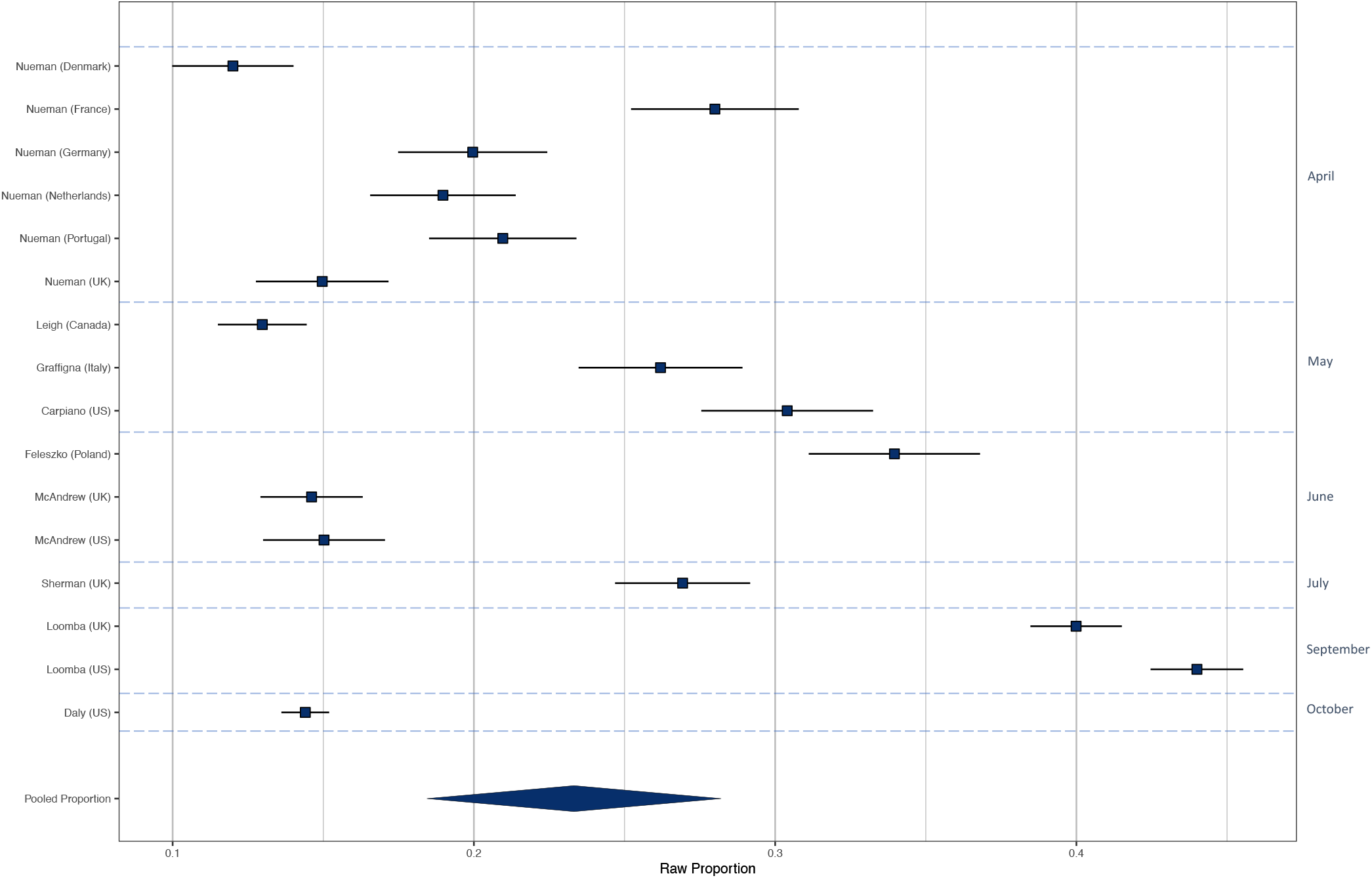
Raw proportions of individuals reporting unsure of vaccination across 16 samples.

**Figure S3.**
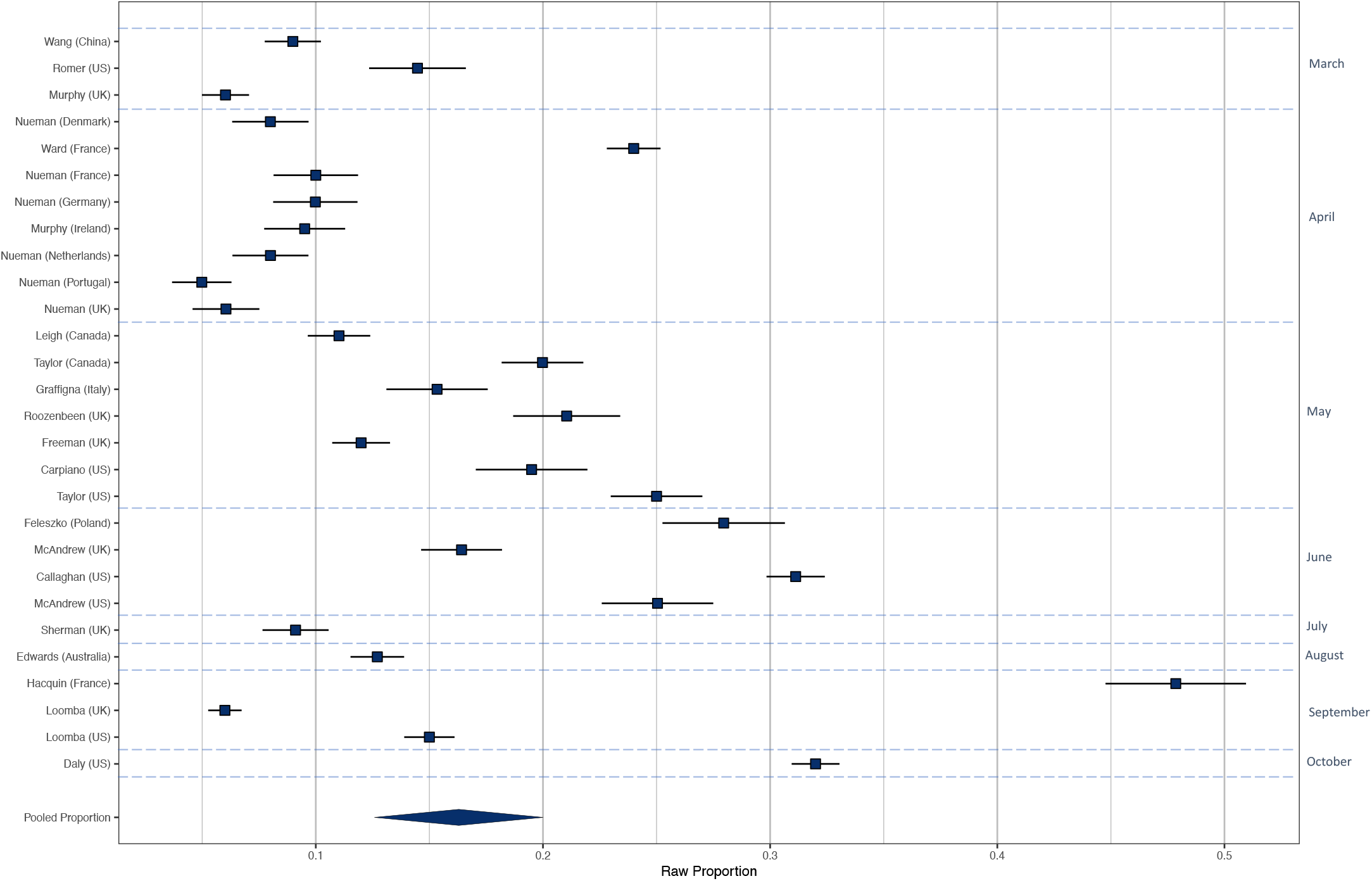
Raw proportions of individuals reporting intending not to vaccinate across 28 studies.

