## Supplementary material for "International estimates of intended uptake and refusal of COVID-19 vaccines: A rapid systematic review and meta-analysis of large nationally representative samples": Online supplementary document

**Online Supplementary Materials**

Publication bias analysis

To examine potential publication bias / small study effects we examined funnel plots for asymmetry using Egger’s regression test. We conducted Trim and Fill analysis to correct for asymmetry around the pooled estimate, which computes i) the number of ‘hypothetical’ effect sizes needed for symmetry and ii) the pooled prevalence with this hypothetical effect sizes included.

Meta-analysis results

***Intending to vaccinate (Yes).*** Including all 28 samples, the pooled proportion of the samples who reported intending to vaccinate was .729 [n = 28, 95% CI: .665 to .784: I2 = 99.6%]. See

Figure S1: Raw proportions of intentions to vaccinate across the 28 samples, additional online material. Leave-one-out analyses demonstrated limited variation in the pooled estimates (min = .715; max = .739). Egger’s test of funnel plot asymmetry was significant (z = 3.69, p < .001). Trim and fill analyses however did not impute any studies. See Figure S4 overleaf.

***Not indenting to vaccinate (No).*** Across the 28 samples the pooled proportion of individuals reporting no intentions to vaccinate was .143 [95% CI: .113 to .178: I^2^ = 99.2%]. See Figure S3: Raw proportions of individuals reporting intending not to vaccinate across 28 studies, additional online material. Leave-one-out analyses demonstrated limited variation in the pooled estimates (min = .135, max = .148). Egger’s test of funnel plot asymmetry was significant (z = -5.28, p < .001). Trim and Fill analyses imputed two studies, which slightly increased the proportion to .153 [95% CI: .121 to .191] . See Figure S5 overleaf.

***Unsure whether to vaccinate (Unsure).*** Across 16 samples the pooled proportion of individuals unsure whether to vaccinate was .220 [95% CI: .178 to .270: I^2^ = 98.9%]. See

Figure S2: Raw proportions of individuals reporting ‘unsure’ of vaccination across 16 samples, additional online material. Leave-one-out analyses demonstrated limited variation in the pooled estimates (min = .209, max = .229). Egger’s test of funnel plot asymmetry was significant (z = -2.63, p = .008). However, Trim and Fill analyses did not impute any studies. See Figure S6 overleaf.

**Figure S4. Intending to vaccinate (Yes) funnel plot.**

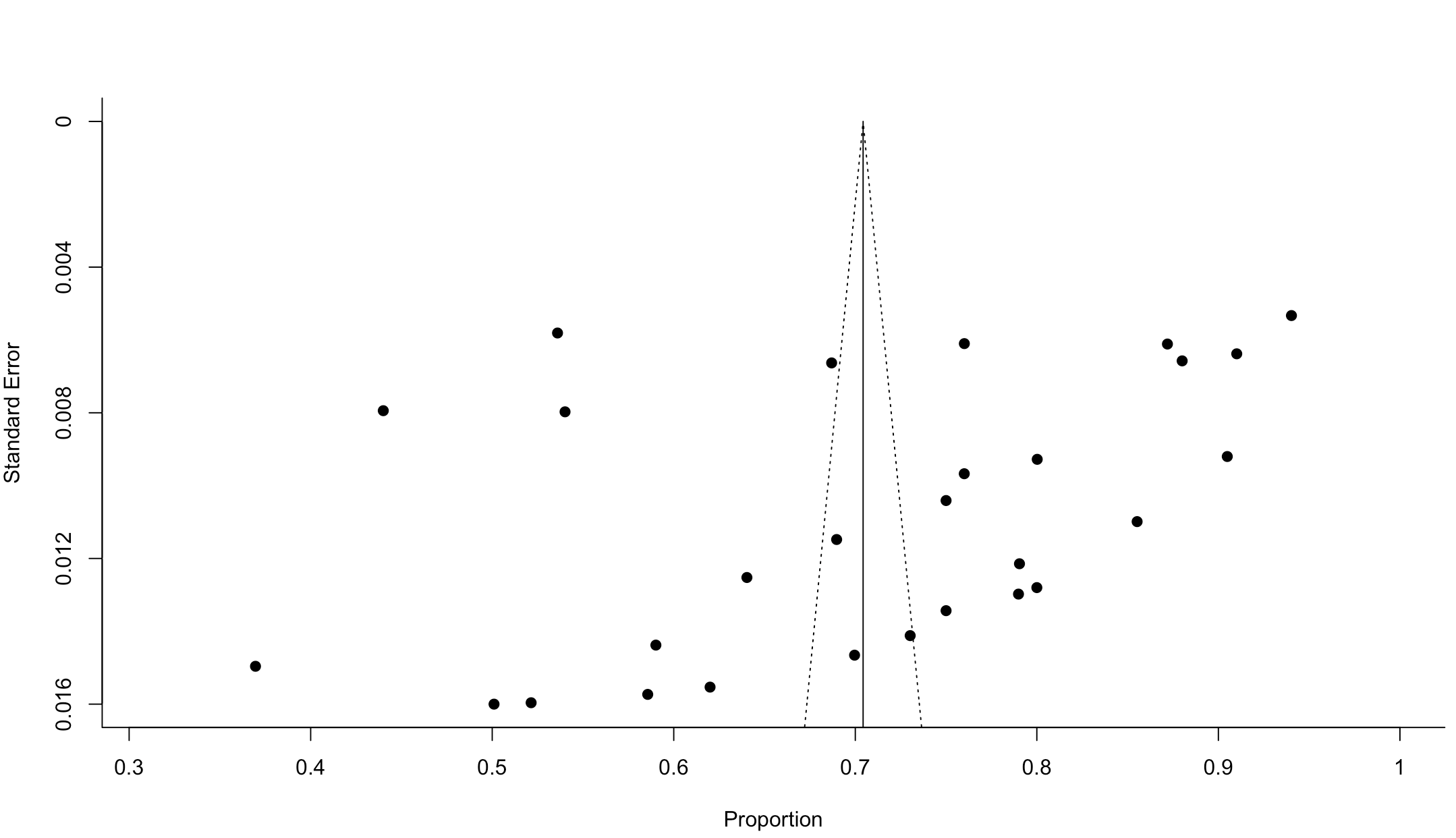

**Figure S5. Intending to vaccinate (No) funnel plot.**

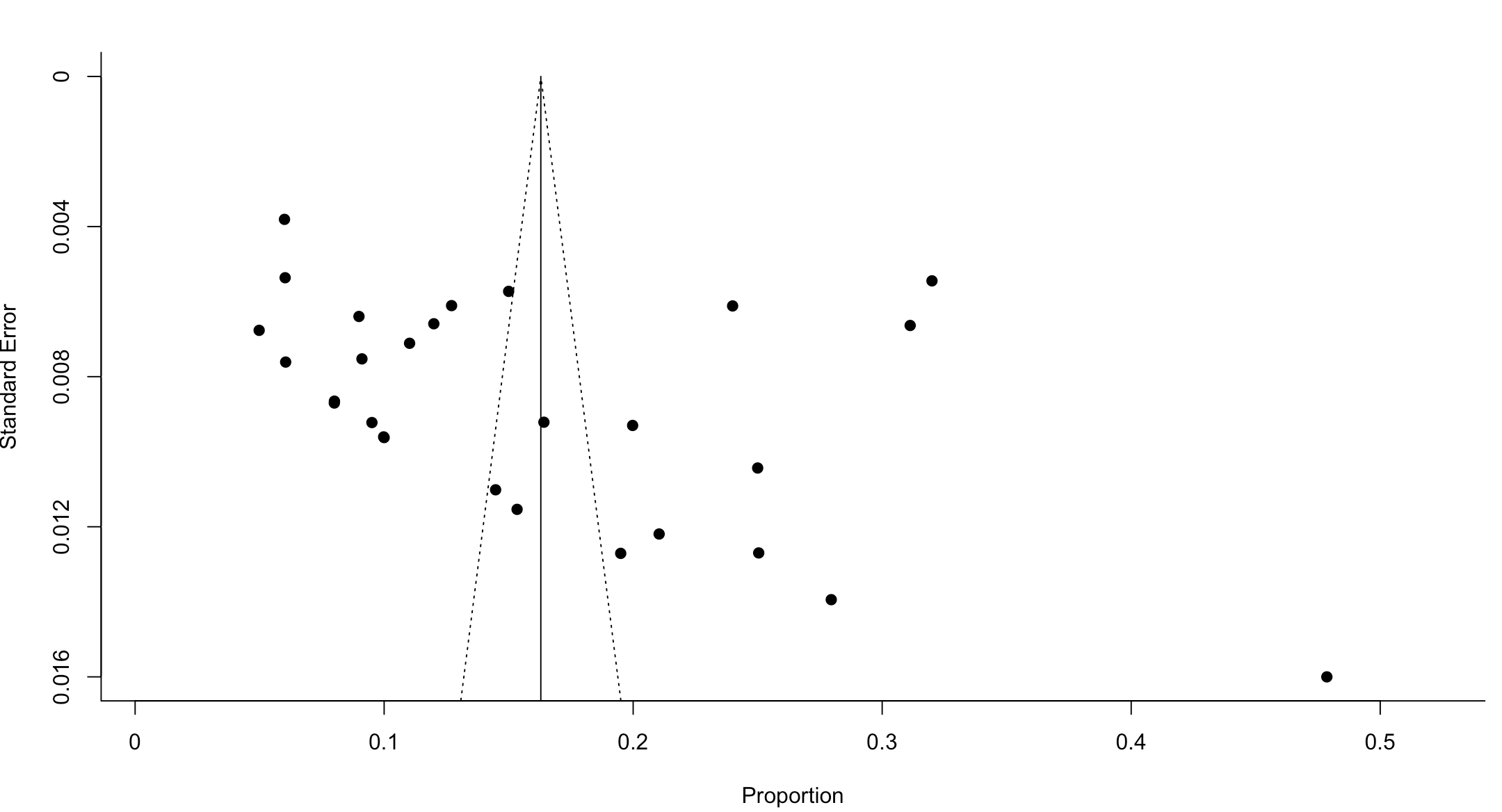

**Figure S6. Intending to vaccinate (Unsure) funnel plot.**

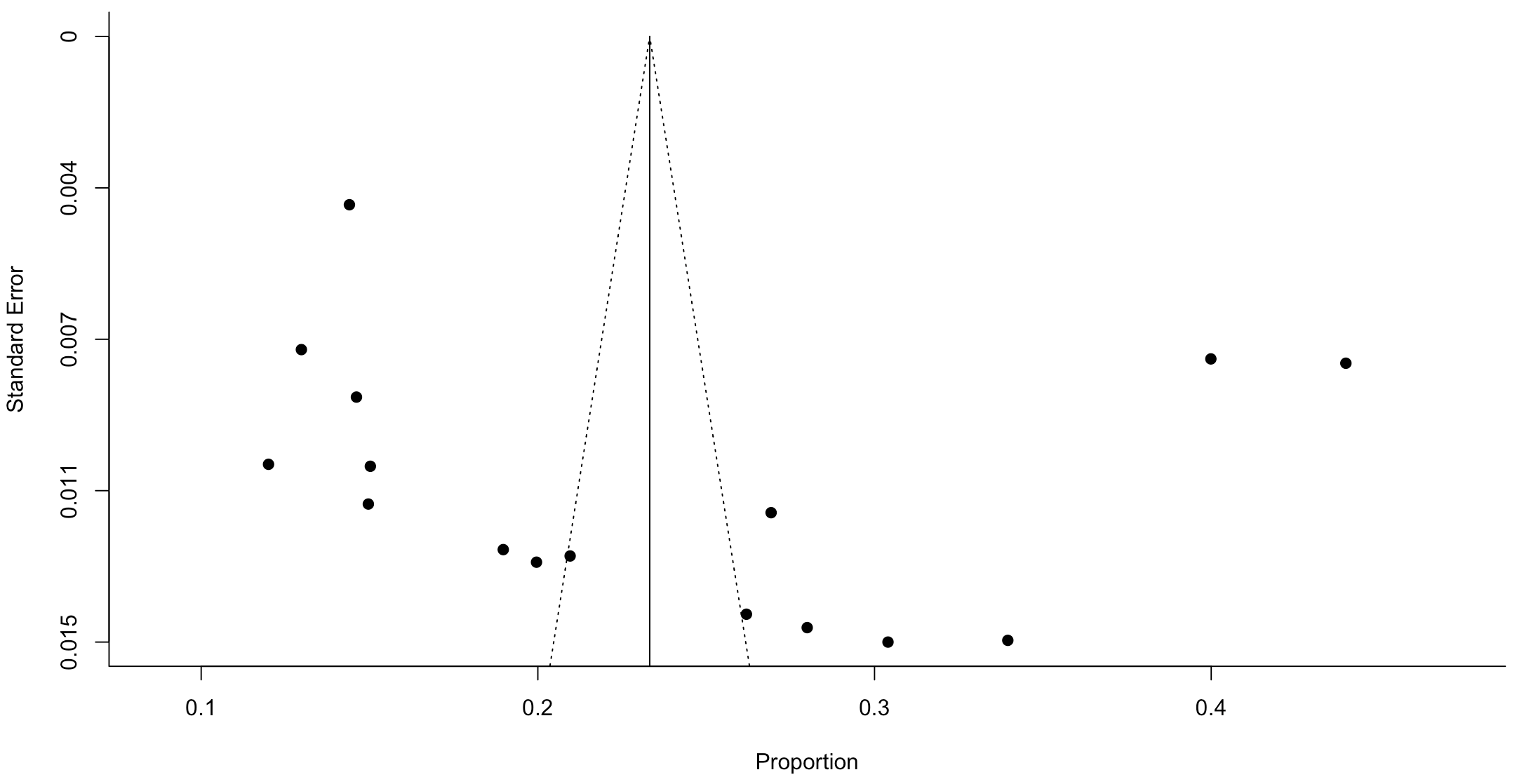

**Sampling method sub-group analysis**. Among studies conducted early in the pandemic there was no significant difference in intentions to vaccinate based on sampling method; quota vs. probability-based sampling (X^2^(1) = 3.02, p = .082). Similarly, there was no significant differences in intentions to not vaccinate (X^2^(1) = 1.60, p = .205). There was a significant difference in unsure responses (X^2^(1) = 4.79, p = .028). In the studies using population sampling the proportion was .282 [95% CI: .243 to .325: I^2^ = 77.0%] and in the studies using quota sampling it was .177 [95% CI: .141 to .220: I^2^ = 95.4%]

***Pre-prints vs journal articles sub-group analysis.*** There was no significant difference in intentions to take a vaccine (X^2^(1) = 1.53, p = .215), not to take a vaccine (X^2^(1) = .005, p = .941), or being unsure (X^2^(1) = 0.01, p = .905) in pre-prints vs accepted articles from data collected early in the pandemic.

List of studies included in review

Callaghan, T., Moghtaderi, A., Lueck, J. A., Hotez, P. J., Strych, U., Dor, A., . . . Motta, M. (2020). Correlates and Disparities of COVID-19 Vaccine Hesitancy. *SSRN pre-print*. https://papers.ssrn.com/sol3/papers.cfm?abstract_id=3667971

Carpiano, R. M. (2020). Demographic differences in US adult intentions to receive a potential coronavirus vaccine and implications for ongoing study. *medRxiv pre-print*. https://www.medrxiv.org/content/10.1101/2020.09.07.20190058v1

Daly M., & Robinson, E. (2020). Willingness to vaccinate against COVID-19 in the US:

Longitudinal evidence from a nationally representative sample of adults from April-October 2020. *medRxiv.. https://www.medrxiv.org/content/10.1101/2020.11.27.20239970v1*

Edwards, B., Biddle, N., Gray, M., & Sollis, K. (2020). COVID-19 vaccine hesitancy and resistance: Correlates in a nationally representative longitudinal survey of the Australian population. *medRxiv preprint*. https://www.medrxiv.org/content/10.1101/2020.11.13.20231480v2

Feleszko, W., Lewulis, P., Czarnecki, A., & Waszkiewicz, P. (2020). Flattening the curve of COVID-19 vaccine rejection—A global overview. *SSRN pre-print*. https://papers.ssrn.com/sol3/papers.cfm?abstract_id=3631972

Freeman, D., Waite, F., Rosebrock, L., Petit, A., Causier, C., East, A., . . . Lambe, S. (2020). Coronavirus Conspiracy Beliefs, Mistrust, and Compliance with Government Guidelines in England. *Psychological Medicine*. doi:10.1017/S0033291720001890

Graffigna, G., Palamenghi, L., Boccia, S., & Barello, S. (2020). Relationship between citizens’ health engagement and intention to take the covid-19 vaccine in italy: A mediation analysis. *Vaccines, 8*(4), 1-11. doi:10.3390/vaccines8040576

Hacquin, A., Altay, S., de Araujo, E., Chevallier, C., & Mercier, H. Sharp rise in vaccine hesitancy in a large and representative sample of the French population: reasons for vaccine hesitancy. *PsyArXiv pre-print*. https://psyarxiv.com/r8h6z/

Leigh, J. P., Fiest, K., Brundin-Mather, R., Plonikoff, K., Soo, A., Sypes, E. E., . . . Fox-Robichaud, A. (2020). A national cross-sectional survey of public perceptions, knowledge, and behaviors during the COVID-19 pandemic. *medRxiv pre-print*. https://www.medrxiv.org/content/10.1101/2020.07.07.20147413v1

Loomba, S., de Figueiredo, A., Piatek, S., de Graaf, K., & Larson, H. J. (2020). Measuring the Impact of Exposure to COVID-19 Vaccine Misinformation on Vaccine Intent in the UK and US. *medRxiv* *pre-print.* https://www.medrxiv.org/content/10.1101/2020.10.22.20217513v1.full.pdf

McAndrew, S., & Allington, D. (2020). Mode and Frequency of Covid-19 Information Updates, Political Values, and Future Covid-19 Vaccine Attitudes. *PsyArXiv pre-print*. https://psyarxiv.com/j7srx/

Murphy, J., Vallières, F., Bentall, R. P., Shevlin, M., McBride, O., Hartman, T. K., . . . Levita, L. (2020). Preparing for a COVID-19 vaccine: Identifying and psychologically profiling those who are vaccine hesitant or resistant in two general population samples. *PsyArXiv pre-print*. https://psyarxiv.com/pev2b/

Neumann-Böhme, S., Varghese, N. E., Sabat, I., Barros, P. P., Brouwer, W., van Exel, J., . . . Stargardt, T. (2020). Once we have it, will we use it? A European survey on willingness to be vaccinated against COVID-19. *Eur J Health Econ, 21*(7), 977-982. doi:10.1007/s10198-020-01208-6

Romer, D., & Jamieson, K. H. (2020). Conspiracy theories as barriers to controlling the spread of COVID-19 in the U.S. *Soc Sci Med, 263*, 113356. doi:10.1016/j.socscimed.2020.113356

Roozenbeek, J., Schneider, C. R., Dryhurst, S., Kerr, J., Freeman, A. L., Recchia, G., . . . van der Linden, S. (2020). Susceptibility to misinformation about COVID-19 around the world. *Royal Society Open Science, 7*(10), 201199.

Sherman, S. M., Smith, L. E., Sim, J., Amlôt, R., Cutts, M., Dasch, H., . . . Sevdalis, N. (2020). COVID-19 vaccination intention in the UK: Results from the COVID-19 Vaccination Acceptability Study (CoVAccS), a nationally representative cross-sectional survey. *medRxiv pre-print*. https://www.medrxiv.org/content/10.1101/2020.08.13.20174045v1

Taylor, S., Landry, C. A., Paluszek, M. M., Groenewoud, R., Rachor, G. S., & Asmundson, G. J. G. (2020). A Proactive Approach for Managing COVID-19: The Importance of Understanding the Motivational Roots of Vaccination Hesitancy for SARS-CoV2. *Frontiers in Psychology, 11*. doi:10.3389/fpsyg.2020.575950

Wang, J., Jing, R., Lai, X., Zhang, H., Lyu, Y., Knoll, M. D., & Fang, H. (2020). Acceptance of covid-19 vaccination during the covid-19 pandemic in china. *Vaccines, 8*(3), 1-14. doi:10.3390/vaccines8030482

Ward, J. K., Alleaume, C., Peretti-Watel, P., Seror, V., Cortaredona, S., Launay, O., . . . Group, C. (2020). The French public's attitudes to a future COVID-19 vaccine: The politicization of a public health issue. *Social Science and Medicine, 265*. doi:10.1016/j.socscimed.2020.113414

Woko, C., Siegel, L., & Hornik, R. (2020). Explaining the association of race and COVID-19 vaccination intentions: the role of behavioral beliefs and trust in COVID-19 information sources. *PsyAXiv pre-print*. https://psyarxiv.com/r3fma/
